# AutopsyPrint: A novel tool for translating ballistic and sharp force injury trajectory findings into 3D printable models

**DOI:** 10.64898/2026.07.10.26357738

**Authors:** Christine E. Parsons, Asser H. Thomsen, Mikkel V. Petersen

**Affiliations:** Interacting Minds Center, Department of Clinical Medicine, Aarhus University, Palle Juul-Jensen Boulevard 99, 8200, Aarhus N, Denmark; Department of Forensic Medicine, Aarhus University, Palle Juul-Jensen Boulevard 99, 8200, Aarhus N, Denmark; Center of Functionally Integrative Neuroscience, Department of Clinical Medicine, Aarhus University, Palle Juul-Jensen Boulevard 99, 8200, Aarhus N, Denmark

**Keywords:** Autopsy, 3D models, Wound trajectories

## Abstract

When autopsy findings are presented on two-dimensional paper-based models, there is an inherent reduction of spatial information, which the viewer must infer from simplified anatomical and geometrical representations. Multiple diagrams representing trajectory angles must be integrated into a complete mental model, introducing potential for errors in viewer understanding. 3D models can address these issues but have shown limited adoption in forensic autopsy reporting given the technical competences and software required to produce them. Here, we present AutopsyPrint, a workflow and open web-based tool for generating 3D printable body models annotated with wound trajectories. The tool supports marking different wound types, including ballistic and stab wounds, on male and female bodies, which can be posed to accommodate a diversity of trajectories. Based on our testing, we present a set of suggested workflow steps and parameters based to facilitate standardization of the 3D models produced, balancing between precision and print time and materials. To ensure accessibility, the tool runs fully in the user’s browser, and all annoted data is stored locally. By making AutopsyPrint open access, we intend to build practical experience with model creation, to ultimately advance the use of 3D models in the field.

---

Accurate, efficient communication of forensic autopsy findings to diverse audiences, including jurors, legal counsel and law enforcement, is a persistent challenge for forensic practitioners (Bolliger & Thali, 2015; Koch, 2025). Autopsy findings are typically recorded using paper-based 2D formats (Garland et al., 2025), a process that entails loss of three-dimensional information. While surface injuries and body markings can be accurately noted on paper models, the recording of wound trajectories poses basic geometric challenges. A single trajectory requires multiple angular components for a complete spatial description. Within autopsy reporting, wound trajectories are often described using three directions: front to back, downward, from right to left (Denton et al., 2006; DiMaio, 2015).To effectively capture these directions on a flat body model, forensic practitioners will often sketch multiple diagrams with the model in front, back, or top-down positions, depending on the injury path.

For the viewer of autopsy findings, their two-dimensional presentation requires inference of spatial information from simplified anatomical and geometrical representations. When multiple diagrams are used to represent wound trajectories, the viewer must also integrate these into a coherent mental model, introducing further potential for inferential error. Forensic specialists have the extensive training, experience and anatomical knowledge required to mentally reconstruct 3D models from 2D diagrams. For jurors and other non-specialist stakeholders, however, such mental reconstruction is likely demanding (Preece et al., 2013), if not impossible. 3D printing technologies can render autopsy findings onto physical models, providing more accessible spatial mapping than afforded by paper-based models (Petersen et al., 2024). Representation of wounds and trajectories on a printed model can allow visual inspection and direct contact with a physical replica, which can be especially useful for communication with different medico-legal stakeholders. The possibility of viewing, manipulating and otherwise interacting with complex autopsy data are key advantages of 3D rendering, and has stimulated the development of Augmented Reality (AR) technologies for mixed-reality digital autopsies (Pooryousef et al., 2024; Pooryousef et al., 2023).

Despite their interactive possibilities and advantages for data capture and report generation, AR technologies have barriers to their development and implementation. These barriers include specialized hardware requirements, software development costs, and user training (Birlo et al., 2022; Juhnke et al., 2025). By contrast, 3D printing of autopsy models can now be achieved with relatively low financial cost and can provide outputs which are easy to understand with minimal training or specialist expertise. The declining costs of fused deposition modeling (FDM) printers, combined with the availability of user-friendly computer-aided design (CAD) software, have created an opportunity for immediate use in forensic practice.

## Communication of findings using 3D printed models

In the broad field of medical data visualization, an increasing number of studies have demonstrated the benefits of 3D models for communication between physician and patient, for medical training, and in multidisciplinary clinical collaborations. For example, 3D models of congenital heart disease have been shown to enhance physician–parents–patient communication in clinical practice (Biglino et al., 2015). A systematic review reported that medical students perceived enhancements to their learning skills and knowledge when 3D printed models were part of their curriculum (Ardila et al., 2023). The use of 3D models has also been shown to enhance critical cardiac care during a simulation training involving multidisciplinary intensive care teams (Olivieri et al., 2016).

In forensic settings, applications of 3D models have ranged from injury reconstruction and weapon comparison to courtroom demonstrations (Carew et al., 2021a; Simon et al., 2022). To date, a major focus has been on the potential influence of model use on the jury, and the archival advantages conferred by printed models (i.e., contributing to long-term documentation). Studies of jury-related responses to printed models have generally suggested that 3D models can be useful aids to comprehension (Blau et al., 2019; Errickson et al., 2020). For long-term documentation, a cited advantage of printed models is that they can be handled without the preservation and safety concerns associated with original, often biological materials (Baier et al., 2018).

## The need for standardizing 3D Printing workflows for Forensic Wound Trajectories

Despite the potential benefits, there has been limited description of the technical process of 3D printing for forensic autopsies, and calls have been made for the development of best practice guidelines (Carew et al., 2021a; Errickson et al., 2022). Currently, there is heterogeneity in 3D printing production practices, with many forensic practitioners developing in-house local procedures, or establishing collaborations with academic partners (Baier et al., 2018; Errickson et al., 2022). The lack of standardization and sharing of best practices represent serious obstacles to the uptake of 3D printing, and for the admissibility of evidence in legal proceedings (Errickson et al., 2022).

To address these obstacles, we developed AutopsyPrint, a novel tool for converting ballistic and sharp force injury trajectory findings into 3D printable models. Our tool is specifically designed to offer a standardized workflow for forensic scientists, who possess extensive anatomical and medico-legal expertise but may have limited experience with 3D rendering and CAD software. We focus on trajectory documentation as it represents one of the most challenging aspects of traditional autopsy reporting to communicate effectively, requiring complex spatial relationships to be interpreted across multiple 2D diagrams.

Here, we detail the development process and functionality of AutopsyPrint. We first consider the main requirements for printing different wound forms (e.g., gunshot, stab), and typical documentation procedures and challenges within standard autopsy workflows. Next, we describe our testing process using different printing parameters, and their implications for the resultant 3D models. From this testing process, we provide a set of guidelines for forensic practitioners wishing to integrate 3D printing into their workflows. Finally, we present a web application, with core digitization and model export functionality, which has the overarching aim of broadening access to 3D model printing, lowering technical barriers to use in real-world practice.

## 2. Methods

### 2.1 Overview

In this article, we describe our development and testing process for an application facilitating 3D printing of body models to document wound trajectories in forensic cases. We aimed to develop a workflow for mapping and exporting findings for 3D printing, augmenting the existing workflow of forensic specialists. Therefore, we first describe the workflow, as well as the specific challenges of data rendering, drawing on forensic expertise from Aarhus University’s Department of Forensic Medicine.

With current 3D printing models, there are a multiplicity of parameters with regards to filament color, body size, and body positioning. Trajectory-related parameters include the method for marking entry and exit into the body (e.g., via a hole, where a cylindrical structure can be inserted post printing) and coloring. Each parameter opens for considerations related to the cost of materials, time to print, model finish, and injury prominence or visibility. We evaluate the different prototypes printed and provide practice points based on our experiences.

#### Materials: hardware and software

All 3D models described here were printed using the Bambu Lab A1 3D printer, combined with the Automatic Material System (AMS) Lite when testing multi-color prints. All models were printed using standard PolyLactic Acid (PLA) filaments. For slicing the exported body models, along with generating and sending print instructions (G-Code) to the printer, we used Bambu Studio software (https://bambulab.com/en/download/studio). Finally, we developed a standalone desktop app, ‘AutopsyPrint’ for Windows 10/11 using the cross-platform game engine Unity (version 6000.0.26f1, https://unity.com/). For demonstration and testing, we built a platform agnostic version of the application using WebGL and deployed it on https://mvpetersen.github.io/autopsyprint/ for open access.

### 2.3 App Requirements

The specific requirements for application functionality identified via forensic workflow analysis are listed in Table 1. Development was focused on streamlining the process of (1) plotting trajectories, (2) posing the model, and (3) directly exporting 3D model file(s) compatible with 3D printing software.

**Table 1.** Identified Application Requirements and a Rationale for Each.

| Requirement | Description | Rationale |
| --- | --- | --- |
| Trajectory types | Map three different types of trajectories: gunshot wound (GSW), stab, and other (e.g., shrapnel). | Allow the user to easily distinguish different types of ballistic injuries (e.g., printed in different colors). |
| Trajectory mapping | Accurately defined entry and, if perforating, exit position and direction. | It should be simple and quick to map trajectory angles from autopsy findings to the digital model. |
| Body pose | Dynamically change the pose of the body model before printing. | Can help avoid trajectories intersecting body parts that they should not (e.g., a GSW to the thorax intersecting with the arms in the default anatomical pose). |
| File export | Export to a file format that supports multiple objects (e.g., body model + trajectories) and is directly compatible with 3D printing software. | Minimize time to print. |
| Model export options | Control settings such as model size, trajectory size, single- or multi-color printing, and additive or subtractive trajectories. | Added end-user flexibility and options for iterating on the design (e.g., printing a quick draft before the final print). |

*Note.* GSW = gunshot wound.

### 2.3 App Development

#### 2.3.1 Trajectory mapping

In previous work (Petersen et al., 2024), we evaluated a tool for digitizing autopsy findings from standard paper formats. The aim of our digital tool was to demonstrate the precision, replicability, and flexibility afforded by interactive visualizations for forensic practice. While digital tools have begun to advance the field of forensic visualization, the challenge of converting complex trajectory data into physical models remains unaddressed. Therefore, the present work develops accessible methods for transforming ballistic and sharp force injury trajectory data to 3D printable models.

To place GSW and stab trajectories, and optionally change the exit trajectory for perforating GSW, we use a spherical coordinate system (elevation and azimuth angle) orientated with a sagittal meridian plane (Figure 1). A trajectory can therefore be defined with an elevation (-90 to 90 degree) angle and an azimuth (-180 to 180 degrees) angle relative to the sagittal and transverse plane. For ease of communication with non-specialists, these coordinates are remapped to describe a trajectory facing the front or back of the model with an azimuth (horizontal) angle pointing 90 degrees left to 90 degrees right and an elevation angle pointing 90 degrees up or down.

**Figure 1:**
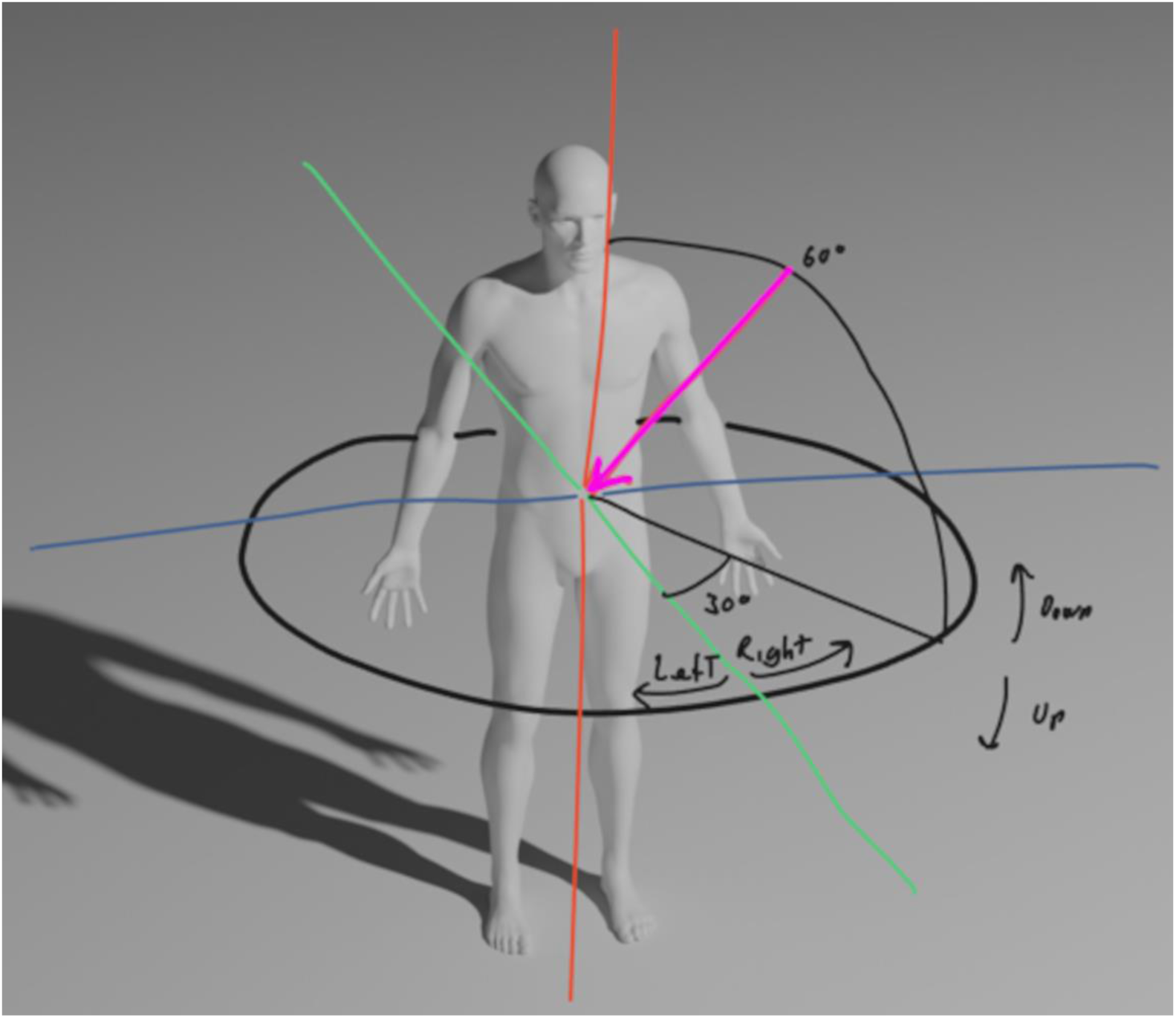
Trajectories are mapped onto the surface of the body using a spherical coordinate system aligned with the vertical axis (polar axis in red), a transverse reference plane (blue and green axes), and a sagittal meridian plane (red and green axes). In this system, trajectory directions are mathematically defined using an elevation angle (-90 to 90 degrees) below and above the reference plane and an azimuth angle (-180 to 180 degrees) relative to the sagittal plane. For ease of communication with non-specialists, coordinates are remapped to describe whether they face the front or back of the body and the direction they *point towards.* The direction of the example trajectory (purple) in the figure is *facing the front, 60 degrees downward, and 30 degrees toward the right*.

#### 2.3.2 3D Body models and rigging

For body models, we obtained a set of male and female body models created by [Andrei Cristea, 2022] (https://lhndo.github.io/undoz/blog.html). We used the freely available Blender 3D modelling software (https://www.blender.org) to rig a virtual skeleton to each of these models (Figure 2). *The developed workflow first requires the models to be rigged in our tool, and then our tool uses inverse kinematics (IK) to dynamically manipulate the skeleton to move the limbs and place the model in different poses*

**Figure 2:**
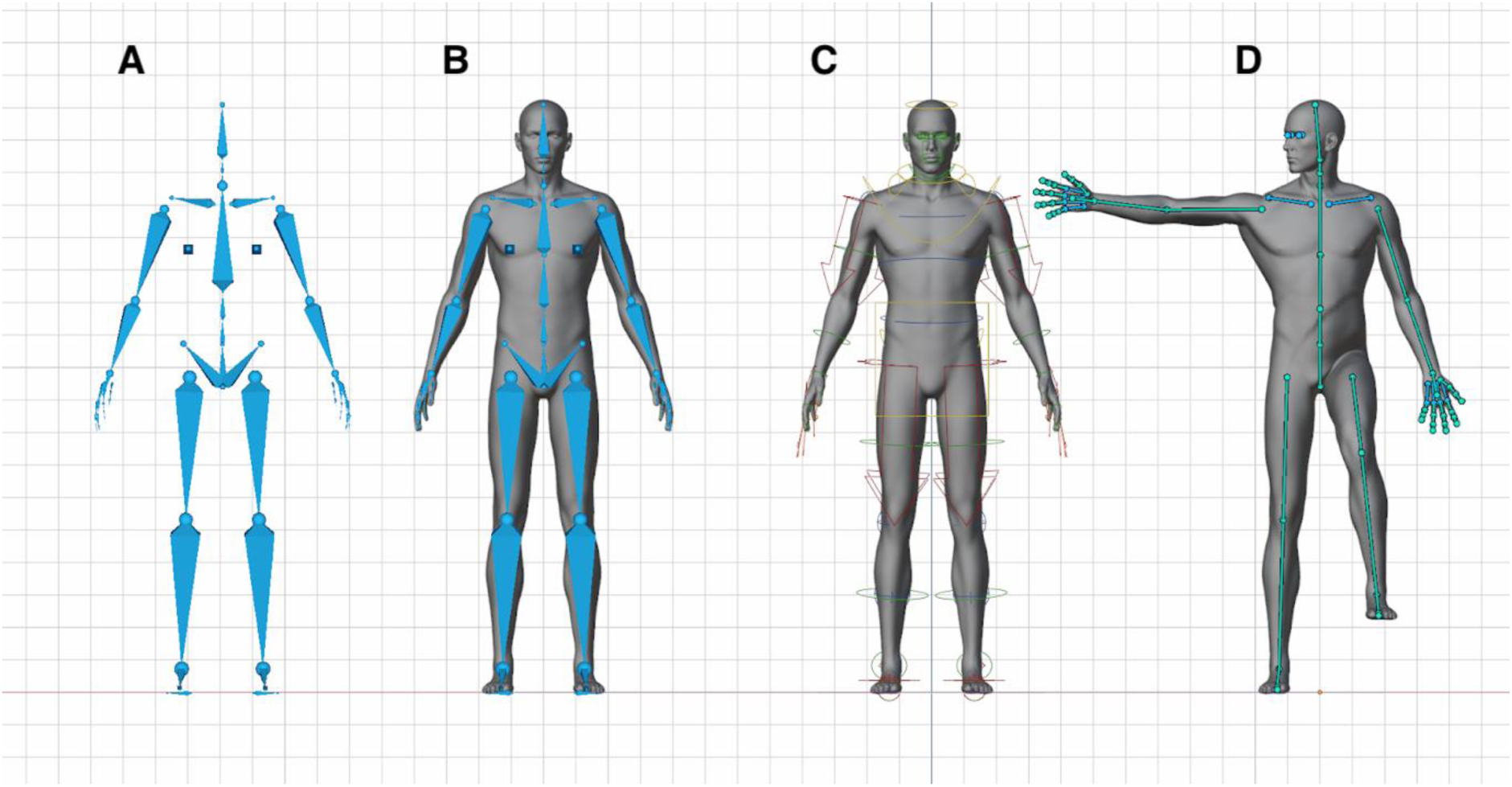
(A-B) A male model rigged with a skeleton using the Rigify add-on in Blender. (C-D) Mesh vertices are automatically grouped and linked to follow individual bones, enabling dynamic manipulation of the model pose.

#### 2.3.3 Pose manipulation in Unity

For each of the four limbs, we pre-defined several pose targets (Figure 3A) to constrain the IK to anatomically realistic positions.

**Figure 3:**
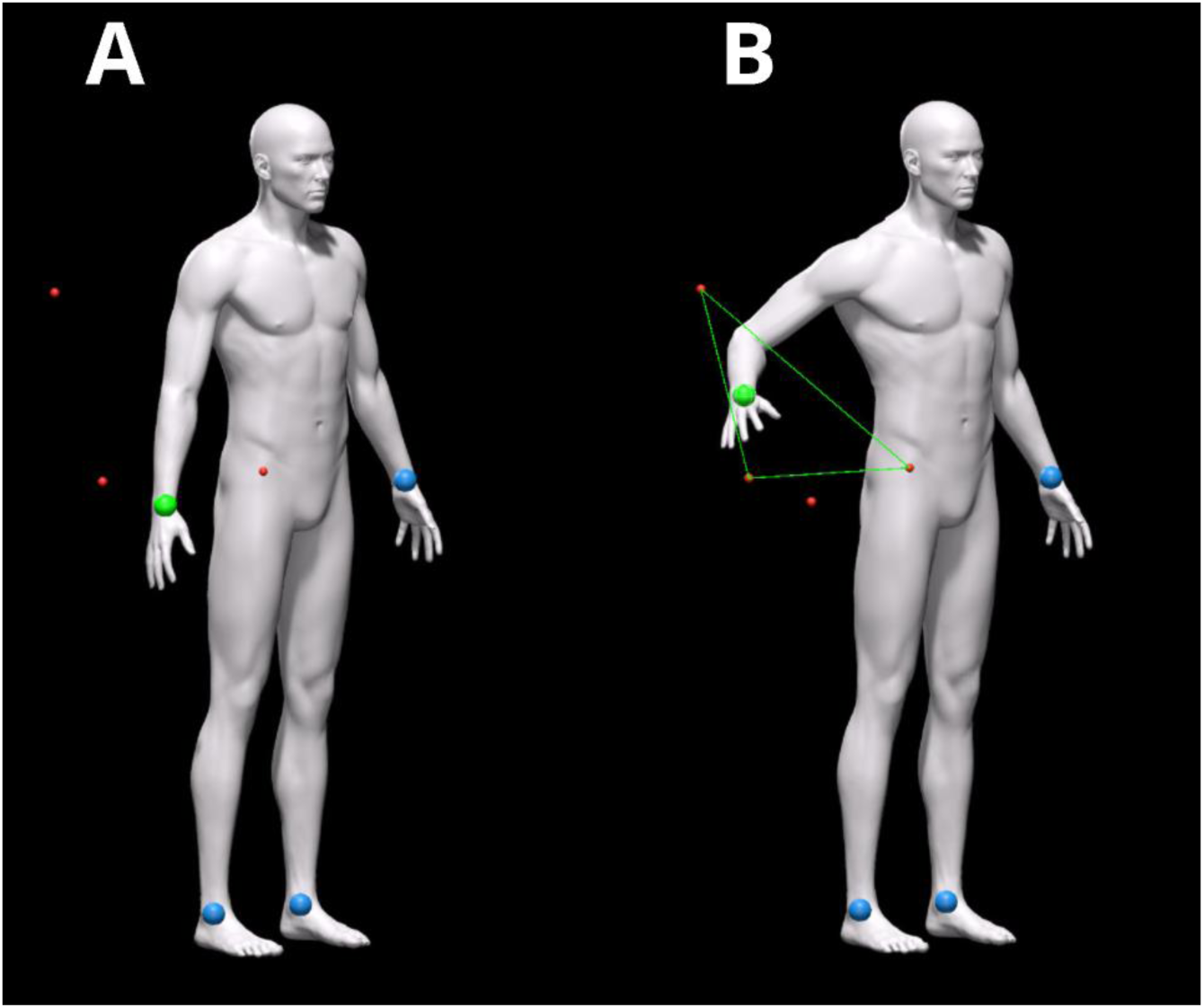
(A) Multiple pose targets are pre-defined for each limb and controlled by an IK-handle that the user can click and drag. (B) The 3D position of the IK-handle (and followed by the associated skeleton bones) is calculated by projecting pose targets to a 2D canvas, identifying the nearest encompassing triangle, calculating the barycentric coordinates and applying these as weights to the 3D position of the pose targets.

To make positioning intuitive, we designed a User Interface (UI) where the predefined pose targets were projected onto a 2D canvas (i.e., screen space). When the user clicked and dragged the IK handle (blue sphere), the 3D position of the associated body part (hand or foot) was interpolated based on proximity to the individual pose target. We projected all pose targets to screen space and identified the nearest triangle of pose targets that the IK handle is inside. Barycentric coordinates are then calculated and used to interpolate between the corresponding world space pose target positions (Figure 3B). If the mouse was dragged outside the triangle area, the IK handle position was projected orthogonally onto the nearest edge or snapped to the nearest pose target.

#### 2.3.4 Trajectory re-positioning

Trajectory mapping was developed to support dynamic model poses. When a trajectory was placed, its position was stored using the triangle index and barycentric coordinates within the intersected mesh triangle. Using these coordinates, instead of a typical world space position, enabled us to recalculate the position after the model has been adjusted to a different pose. The trajectory spatial orientation was stored using quaternions and calculated relative to the local coordinate frame defined by the normal and tangent vectors at the specified entry coordinate. This enables robust recalculation of trajectory orientation following body model pose changes.

#### 2.3.5 3D printing model options

We implemented several optional settings for exporting models for printing. Model size could be selected across three standardized sizes: small, medium, and large models. Model color could be customized both for the base model and trajectory types, allowing alignment with available printing filaments or user preferences. The default color scheme assigned white filament to the base anatomical model, with wound paths differentiated by injury type: red for gunshot wounds, blue for stab wounds, and green for other types. The colors can be manually overwritten e.g. for splitting a single injury type into multiple groups by color.

Two options were included for trajectories (1) additive, printing the full trajectory or (2) subtractive, printing a hole corresponding to a trajectory cylinder. For simplicity, the hole diameter was set to 1.8mm, allowing the user to insert a portion of print filament and place it in the hole. For additive trajectory printing, the system was developed to support both multi-color and single-color outputs, accommodating varying 3D printing setups.

#### 2.3.6 File export (.3mf)

By using the .3mf file format, we embedded data for multiple separate parts (body model + trajectories), as well as specifying the filament each part should be printed with. This assumes that the specified number of filaments are available in the 3D printing system. Additionally, it is possible to have parts designated as ‘negative’, subtracting their shape from the base model (e.g., have the trajectory create a hole in the base model). In this work, we adapted the .3mf format for use with Bambu Studio, which employs the 3MF Production Extension. Additional work may be required to ensure compatibility with systems that only support the 3MF Core Specification.

### 2.4 Web application

We deployed a version of the application to a cloud server to enable access to the tool without installing software on local devices. The web application was designed to minimize the learning curve for new users, who would like to augment their current workflows and explore AutopsyPrint.

## 3. Results

### 3.1 Workflow

The overall workflow is demonstrated in Supplementary Video 1 (https://mvpetersen.github.io/autopsyprint/video). In brief, the workflow comprised of: placing trajectories, posing the model, selecting 3D print settings and exporting to the .3mf format. Exported .3mf files can then be imported and printed using Bambu Studio software.

### 3.2 User Interface

Three different modes were implemented (1) Annotation mode, where the model was locked to the default anatomical pose for placement of entry and exit trajectories, (2) Pose mode, where the user could dynamically adjust the model pose, and finally (3) Print mode, where the posed model and trajectories could be exported to a 3D print compatible file format.

### 3.3 Example Autopsy Cases

We created three example autopsy cases showcasing model poses for different entry and exit trajectories (Figure 4).

**Figure 4:**
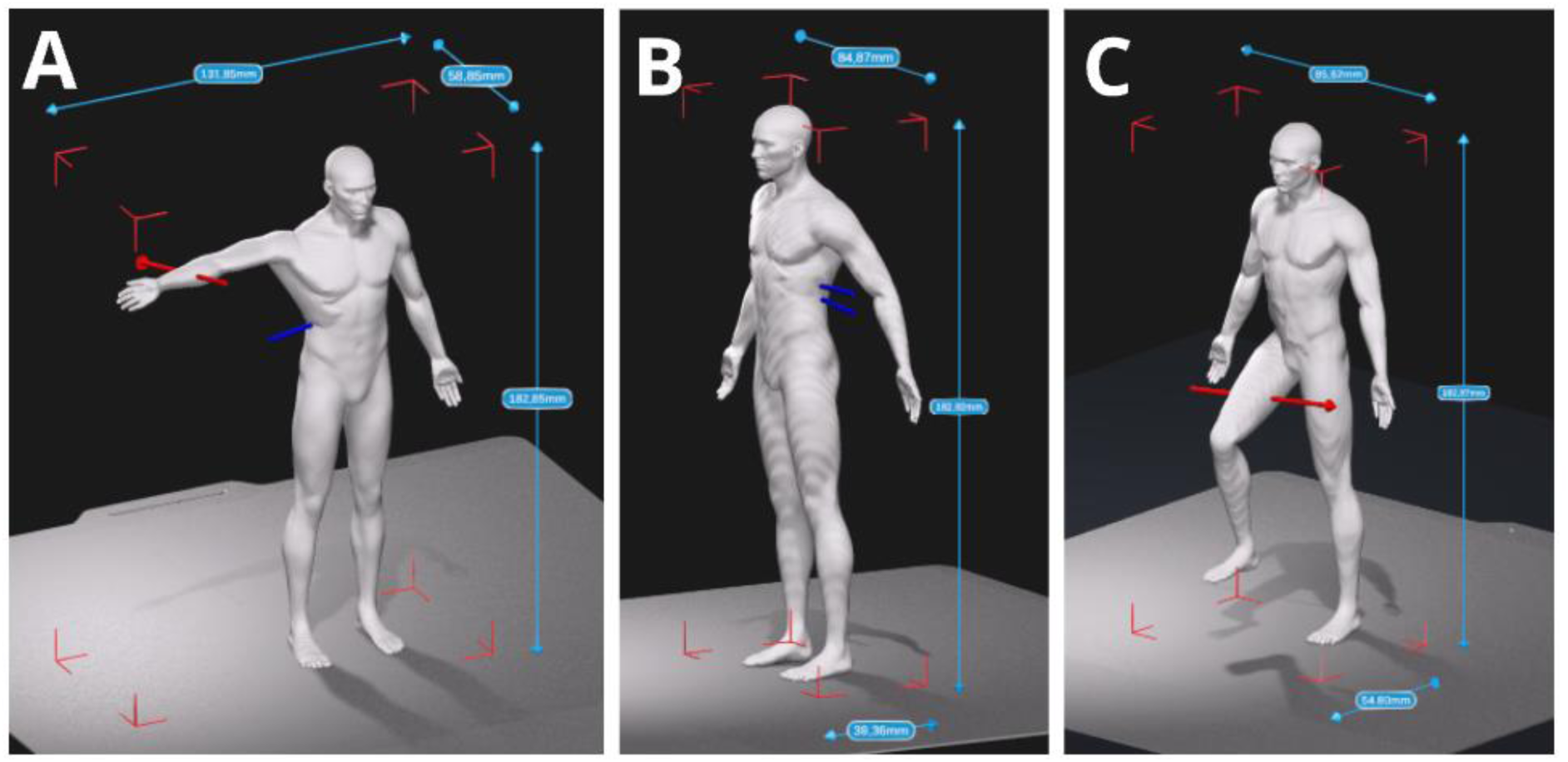
(A-C) Three different example autopsy cases necessitating different model poses for unimpeded visualization.

### 3.4 3D Print Evaluation

Using the three autopsy cases, we exported varying model size, trajectory type and size and print settings (layer height). Table 2 summarizes the print duration and filament use (model and support material) for the different model and print configurations tested. All models were printed with auto-generated tree support (Figure 5B).

**Figure 5:**
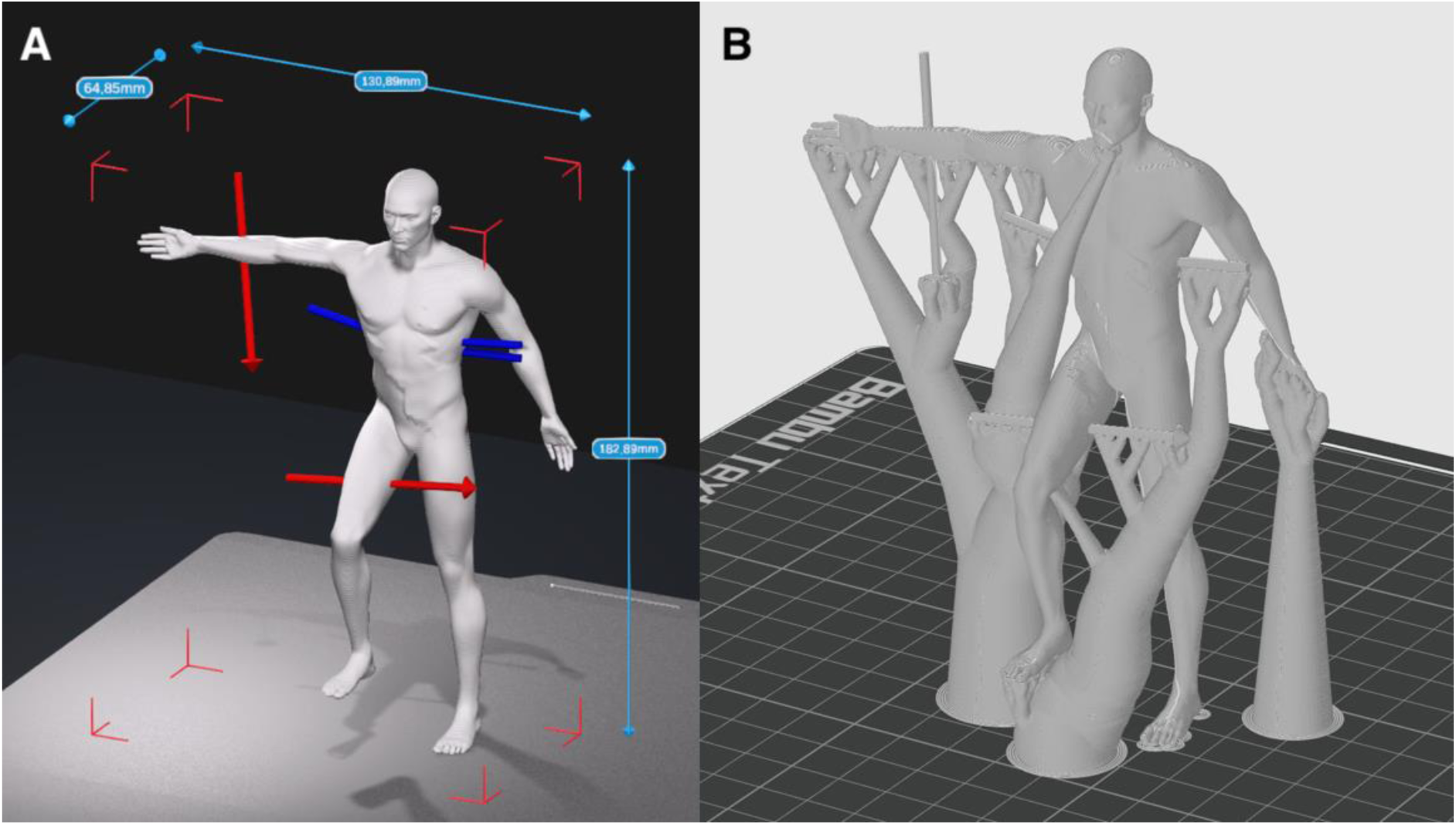
Evaluating the impact of trajectory orientation and model poses when exporting and generating 3D slicing code (A) We created a composite model of the three sample cases presented in Figure 4. (B) We used the slicing software (Bambu Studio) to auto-generate support structures (using the Tree type) for printing free floating parts of the model.

**Table 2.** Overview of Relative Print Duration and Filament Use with Different Model Configurations Using the Combined Autopsy Case.

| Model configuration |  |  | 3D print<br>Layer height | Print<br>duration | Filament |  |
| --- | --- | --- | --- | --- | --- | --- |
| Trajectory | Size | Colors |  |  | Model | Other* |
| Subtractive | S | Single | 0.28mm (extra draft) | 57m | 12g | 7g |

| Model configuration |  |  | 3D print | Print | Filament |  |
| --- | --- | --- | --- | --- | --- | --- |
| Trajectory | Size | Colors | Layer height | duration | Model | Other* |
| Additive | S | Single | 0.20mm (std) | 1h21m | 13g | 9g |
| Additive | S | Single | 0.08mm (high quality) | 3h32m | 13g | 9g |
| Additive | S | Multiple | 0.20mm (std) | 7h15m | 13g | 103g |
| Subtractive | M | Single | 0.28mm (extra draft) | 1h44m | 30g | 19g |
| Additive | M | Single | 0.20mm (std) | 2h23m | 30g | 19g |
| Additive | M | Single | 0.08mm (high quality) | 6h37m | 31g | 21g |
| Additive | M | Multiple | 0.20mm (std) | 10h26m | 30g | 148g |
| Subtractive | L | Single | 0.28mm (extra draft) | 2h54m | 57g | 92g |
| Additive | L | Single | 0.20mm (std) | 4h2m | 57g | 41g |
| Additive | L | Single | 0.08mm (high quality) | 10h56m | 58g | 35g |
| Additive | L | Multiple | 0.20mm (std) | 14h17m | 57g | 204g |
| Additive | L | Multiple | 0.08mm (high quality) | 38h56m | 58g | 423g |
| Additive | L | Multiple | 0.08mm (high quality),<br>0.2mm nozzle | 65h54m | 56g | 324g |

*Note.* The selected 3D printer was the entry-level Bambu Lab A1 with a standard 0.4mm nozzle. All model slicing used the default autogenerated tree support (build plate only). Print duration is the model printing time excluding any preceding calibrations. Model slicing and the associated print duration and estimated filament use were generated using Bambu Studio version 2.6.0.51. The actual printing time of the generated models will depend on the number of trajectories and the model pose.
\* Other filament usage covers support material (model support, purging tower, and filament purging).

#### 3.4.1 Model Size

We evaluated three standardized model sizes (small: 13cm, medium: 18cm, and large: 24cm). The medium scale (18cm) provided a balance between detail preservation, efficiency of printing, and material use. While the large 24 cm models offered increased visibility of trajectory paths, they required substantially longer print times (approximately 2 - 2.5 times longer than the small version) and more material. Small models, though efficient to produce, had more compromised trajectory representations. Another issue with the smaller models was related to printing practicalities, where it became challenging to remove support materials without affecting the model (e.g., finer details like fingers or trajectories can break off and the surface finish of the model can be negatively impacted).

#### 3.4.2 Trajectory type (additive vs. subtractive)

We compared additive and subtractive approaches for trajectory reconstruction. In the additive method, trajectories were printed as cylinders extending from the entry points, while the subtractive method printed small holes in the body models. The additive approach required a single model export but also required additional support structures for angles exceeding 45 degrees from vertical. Subtractive trajectories did not need additional support and allowed for post-print insertion of filaments to visualize different wound trajectories.

#### 3.4.3 Printed layer height

Layer height significantly impacted both model resolution and printing efficiency. Testing across common layer heights (0.08mm, 0.2mm, and 0.28mm), we found that 0.2mm provided a balance between detail retention and print time. While 0.08mm layers offered marginally improved trajectory surface finish, they increased print times by approximately 250-300% compared to 0.2mm layers, without providing meaningful improvements in trajectory detail. The 0.28mm setting, though faster, showed visible stepping on trajectory paths that could impact evidence presentation.

### 3.5 Web application

To allow convenient access for forensic scientists to test AutopsyPrint, we developed and deployed a web application using WebGL (https://mvpetersen.github.io/autopsyprint/). The application enables users to create and export custom autopsy cases for testing a 3D printing workflow (Figure 7).

**Figure 6:**
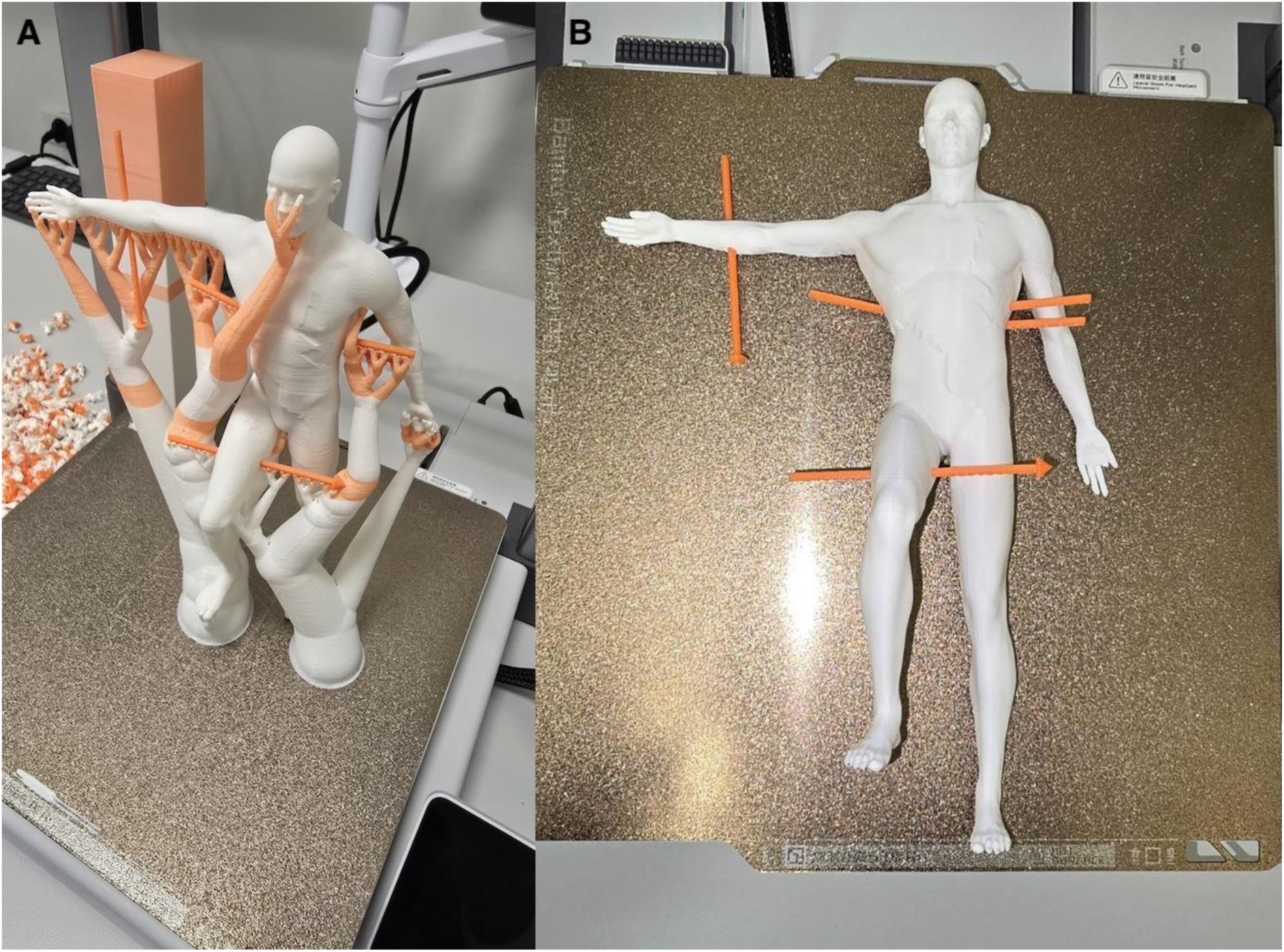
(A) Model of combined autopsy case printed in size Large. (B) Support material should be carefully removed when interfacing with trajectories, to avoid breaking them.

**Figure 7:**
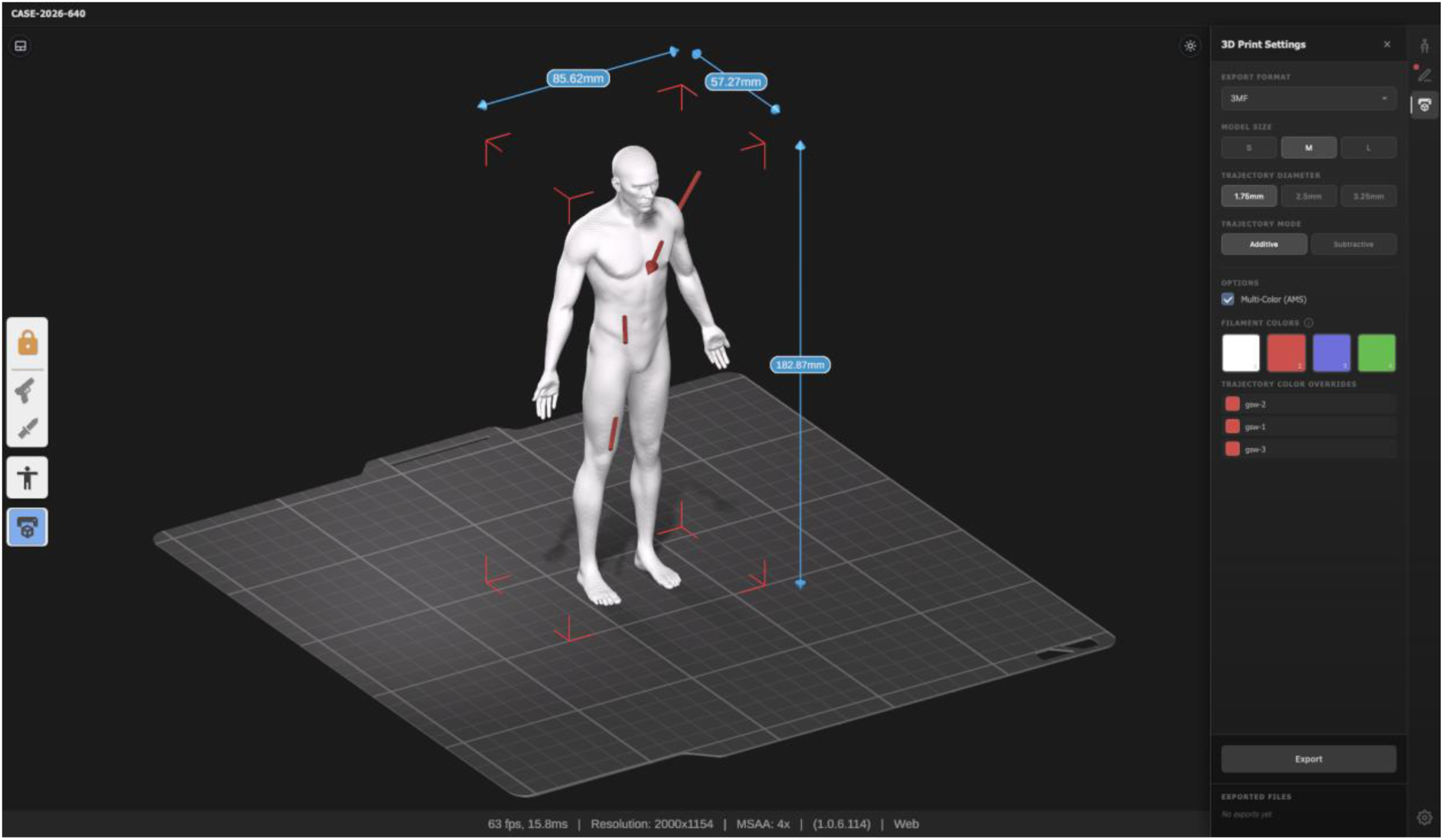
A web application along with illustrative example cases is available on https://mvpetersen.github.io/autopsyprint

## Discussion

In this study, we present a novel application for forensic scientists to translate ballistic and sharp force injury trajectory findings into 3D printable models, with the aim of enhancing autopsy evidence presentation. In a forensic setting, the benefits of 3D printing are manifest. The forensic specialist can mark and review the trajectory findings in a matter of minutes. The body models can be printed within several hours, at a layer height of 0.2 mm and at a material cost of less than $3/unit. Multiple prototypes of the body models can be tested, the digital models modified in response to practitioner observations, and revised body models reprinted within a few hours.

Prior work has clearly demonstrated the value of 3D physical models (e.g., skulls) for communicating information to jurors and stakeholders who do not have extensive medical or technical training (Carew et al., 2021a). Our work focuses specifically on communicating wound trajectory or directional information, an application for which 3D models seem particularly well-suited, and whose viability has already been indicated in other studies (Carew et al., 2021b).

We consider the replicable technical workflow presented here, including tested printing parameters (Table 2), as an answer to calls for greater consistency and standardization in forensic 3D printing practice (Errickson et al., 2022; Srivastava et al., 2025). To this end, we provide an open, accessible web application for interactive 3D mapping of trajectory data and its export to two standard 3D printing formats, .3mf and .stl. Our aim is to allow clinical specialists to test printed body models in their own workflows, minimizing barriers to printing in real world practices. 3D printed models may be used, for example, in teaching, training and in communication with police and within other stakeholder collaborative contexts to enhance understanding of complex spatial relationships (Jani et al., 2021). As discussed elsewhere, a major factor limiting 3D printing’s widespread implementation is a lack of awareness of its applications (Fullerton et al., 2014), limited access to software and training (Jani et al., 2021), as well as an initial learning curve in creating models. Our work addresses these factors.

### Lessons for forensic practitioners: a suggested workflow and parameter selection

From our testing of different 3D printing workflows, we provide a set of suggested practices to streamline implementation. Our first suggested step is to print an initial small model using a large layer height and a single color. This model can be used to iterate and adjust printing parameters, based on how effectively it communicates trajectory findings. For example, the direction of trajectories, relative to the printing plane and model surface can impact the small model print results, so a further iteration may be needed. The user can subsequently adjust the configuration or size, as required (see Table 2), and print the final multi-color model, if available, using the finest necessary layer height. An initial prototype can, from our experience, allow the user to refine autopsy finding presentation before committing to a longer print time for a final model.

For layer height, there is a trade-off between speed of printing and printed detail. At coarser layer heights settings, printed models will have visible layer lines. Layer lines will be especially pronounced for small structures, such as cylindrical trajectories. Finer layer heights will always produce smoother surfaces. We printed all our test models using the standard nozzle of 0.4mm. The minimum recommended layer height (from the manufacturer) for this nozzle size is 0.08mm. Using a 0.2mm nozzle, it is possible to print layer heights down to 0.06mm, resulting in an improved printed model finish, but at a cost of increased print duration. From our examination of models presenting gunshot, knife or other ballistic wounds, we suggest that a layer height of 0.2mm is an acceptable compromise between detail and printing time for medium and large 3D models. For small models, we suggest using the smallest available layer height.

## Limitations

We note several limitations to our rendering design in AutopsyPrint. First, posing the body model created visible artifacts, specifically creases, on parts of the trunk of the model (visible in Figure 6). While such creases are not anatomically realistic, we judged them to be sufficiently minor as not to compromise the demonstrative value of the printed models. As discussed elsewhere, models will always differ from the object they are meant to represent but should meet the requirement of being accurate resemblance of the relevant data (Baier et al., 2018). Furthermore, there is a balance to be achieved with realism of 3D body models, and accurate capture of spatial relationships. Indeed, recent work has shown that realistic models can distress mock jurors (Fawcett et al., 2025), suggesting that partial deviation from anatomical accuracy may be reasonable.

To minimize artifacts, the body model can be further refined via rigging and adjusting the weight painting, which controls how mesh vertices are linked to individual bones. However, such rigging and adjustment would require expertise in 3D artistry, beyond the scope of standard forensic science.

Users may also wish for greater flexibility when posing the model, which is currently constrained within a range of predefined limb positions. Offering less constrained posing would require a more complex user interface and would make it harder to prevent anatomically impossible body model configurations.

Other limitations are technical and print-specific in nature. Printing holes for post-print insertion of filament can be challenging depending on the angle required, relative to the horizontal print plane. The printed hole may not have the correct diameter, meaning it is necessary to manually adjust the diameter post printing, using a small hand drill. Finally, smaller and delicate body areas, such as fingers, can break off when removing support material.

## Conclusion

3D printing offers flexible, inexpensive manufacturing of body models, which can be used for forensic documentation, as well as a demonstrative aid for autopsy case presentation. A major contribution of our work is in sharing a method for printing of forensic cases that is accessible and replicable. Such a tool may be of particular value in resource-limited settings, or in small centers where there is limited capacity to develop in-house workflows (Srivastava et al., 2025). Forensic practitioners can digitize their paper-based autopsy findings using our online web application and export to a 3D printing format within a few minutes. A physical model can then be produced within hours, depending on model size and other print parameters. Capacity is limited only by access to a relatively low-cost 3D printer.

## Data Availability

Example data is available on https://mvpetersen.github.io/autopsyprint.

https://mvpetersen.github.io/autopsyprint/video

